# Study of the Effect of Intestinal Microbes on Obesity: A Bibliometric Analysis

**DOI:** 10.1101/2023.04.01.23288035

**Authors:** Zehao Su, Chenyu Tian, Guan Wang, Jingjing Guo, Xiaoyan Yang

## Abstract

**Background:** Obesity is a serious public health problem. According to statistics, there are millions obese people worldwide. The intestinal microbiota is the microbiota that lives in the gastrointestinal tract of the human body and can interact with the external environment through diet. Probing and summarizing the relationship between intestinal microbes and obesity has important guiding significance for the accurate control of the research direction and expanding the choice of obesity treatment methods. We aimed to use bibliometrics to qualitatively and quantitatively analyze the published literature and to reveal the research hotspots and development trends in the effects of intestinal microbes on obesity research.

**Methods:** We obtained documents from the core collection of Web of Science (WoSCC) from 2013–2022 on February 2nd, 2023. Microsoft Excel, Biblioshiny R language software packages and CiteSpace were employed to collect publication data, analyze publication trends, and visualize relevant results.

**Results:** We identified 8888 original research articles on the effects of intestinal microbes on obesity published between 2013 and 2022. China made the highest number of publications on the topic, with 3036 publications (34.16%), and the United States with 87065 citations made the greatest contribution. University of Copenhagen was the most prolific institution (209 publications). Patrice D. Cani published the most related articles (97 publications), whereas Peter J. Turnbaugh was cited the most frequently (3307 citations). Nutrients was the journal with the most studies (399 publications, 4.49%), and Nature was the most commonly co-cited journal (6446 citations). The burst keywords “obese patient” and “serum” recently appeared as research frontiers.

**Conclusion:** Although China has the largest number of publications, the United States, depending on its largest citations, has become the leader in the effects of intestinal microbes on obesity research. Current research hotspots focus on related mechanisms of the effects of intestinal microbes on obesity and therapeutic methods for obesity with intestinal microbes. Through diet, probiotic preparations and regulation of the intestinal flora, fecal microbe transplantation may have a significant impact on reducing the obesity epidemic. Considering the great potential and application prospects, intestinal microbe applications in obesity therapy will remain a research hotspot in the future.

## 1 Introduction

Obesity is a serious public health problem. According to statistics, there are approximately 600 million obese people and 1.9 billion overweight individuals worldwide (El-Sayed Moustafa and Froguel, 2013). It is predicted that the number of obese people in the world will reach 1.12 billion by 2030 (KellyYang et al., 2008). As far as we know, obesity is not only a change in appearance but also associated with disorders of lipid and glucose metabolism, chronic inflammation, oxidative stress and increased risk of a variety of diseases. According to research, obesity is one of the nonnegligible reasons for cardiovascular disease, diabetes and malignant tumors (BoccellinoDi Domenico et al., 2018; Scherer and Hill, 2016), and it is estimated that more than 80% of the global burden of diabetes and cardiovascular disease will occur in developing countries such as China and India in 2025 (MohanGupta et al., 2016). However, the etiology of obesity is not yet completely clear. The increasing prevalence of obesity will not be explained by changes in energy balance alone. Therefore, it is vital for the whole world to determine the formation mechanism of obesity and then explore effective methods to treat it and reduce the incidence and mortality of obesity-related diseases.

The intestinal microbiota is a microbiota that lives in the gastrointestinal tract of the human body and can interact with the external environment through diet (AlessandroMarcello et al., 2017). These microbes colonize the intestinal tract through mother-to-child transmission from the prenatal period. The colonization of human intestinal microbes continues after birth and is regulated by factors including gestational age, mode of delivery (natural delivery or cesarean section), diet (breastfeeding or baby food), hygiene and antibiotics. The environment and diet of the first three years of life are essential for the acquisition of adult-like microbiota and the establishment of bacteria-host symbiosis that affect immune and nervous system development. At the age from 2 to 5, the intestinal microbiota will gradually stabilize, and there is no significant difference with intestinal microbiota in adults (RodríguezMurphy et al., 2015). Under normal circumstances, the intestinal microbiota maintains a symbiotic relationship with the human body, and a good symbiotic relationship between them is very important for the maintenance of human health (JandhyalaTalukdar et al., 2015), but instantaneous changes in the intestinal ecosystem can lead to the destruction of the symbiotic relationship between microorganisms and hosts, such as aging, obesity, sedentary lifestyles, and dietary patterns, and the effects of antibiotics can change the intestinal microbiota (NazliYang et al., 2004). Because of the important role of the intestinal ecosystem in maintaining host physiology, its changes can also cause a variety of physiological disorders in the human body, including low-grade inflammation, metabolic disorders, excessive lipid accumulation and loss of insulin sensitivity, in turn leading to changes in the composition of the intestinal microbiota and decreasing the diversity of flora and metabolic pathways (Halmos and Suba, 2016; Sidhu and van der Poorten, 2017), thus increasing the risk of metabolic diseases such as obesity and diabetes (TangKitai et al., 2017). Probing and summarizing the relationship between intestinal microbes and obesity has important guiding significance for the accurate control of the research direction and expanding the choice of obesity treatment methods.

Bibliometrics is a discipline that uses mathematics and statistics to quantitatively analyze literature and information. Through quantitative analysis of the subject literature, we can construct its knowledge structure and explore its development trend. At present, it has been widely used in many research fields (GaoDou et al., 2021; LiuGao et al., 2022; WangMa et al., 2020). Overall, this analysis method and the traditional literature review are both based on previous research, summarize the current situation and shortcomings of the research, and finally guide further research. Literature review emphasizes the content of the article, that is, to summarize the aspects and shortcomings of the existing research, and most of them quote the representative papers in the literature according to a preset research context. On the other hand, bibliometric analysis places more emphasis on the analysis of “quantity”. There is no need to analyze the research content of each document in detail but to analyze the number of published documents, the distribution of authors and the relationship of citations in the target field. Most of the included documents are highly quoted. Compared with the traditional literature review, bibliometric analysis can make a more intuitive systematic analysis of all the literature in this field in a visual way, which is helpful for researchers entering a new field to grasp the overall trend of the field. There have been many review articles in the field of intestinal microbes and obesity, but the quantitative analysis in the field is insufficient. Therefore, in addition to briefly summarizing the hotpots in review articles, we use bibliometrics to qualitatively and quantitatively analyze the published literature and intends to reveal the research hotspots and development trends in this field.

## 2 Materials and methods

### 2.1 Data sources and search strategies

Data were obtained from the core collection of Web of Science (WoSCC), a database of Clarivate Analytics. The following search strategies were presented: ((overweight OR obesity OR obes* OR over$weight) AND (gastrointestin* OR gastro-intestin* OR gut OR intestin*) AND (bacteria OR prebiotic OR probiotic OR microbiot* OR microbiome* OR flora OR microflora)). In the end, a total of 8888 original articles in English were retrieved. All the documents were downloaded as “Full Record and Cited References” and saved as “Bibtex” and “plain text file”. The above data were obtained on 2nd February 2023.

### 2.2 Data analysis and graph acquisition

The corresponding files retrieved from WoSCC were imported into Microsoft Excel (16.66.1), Biblioshiny and CiteSpace (6.1. R6) to perform a bibliometric and visual analysis. Annual publications, country annual publications, and publisher annual publications were analyzed with Microsoft Excel. The Biblioshiny platform provides a web-based graphical interface in the Bibliometrix package, which was used to conduct total citation, average article cited, publications of journal and country collaboration map. CiteSpace is interactive visualization analysis software developed by Professor Chen Chaomei to conduct different types of network analyses, such as country collaboration networks, institution collaboration networks, author collaboration networks, co-cited author collaboration networks, co-cited journal collaboration networks, and keyword co-occurrence, which aid in visually analyzing the knowledge domain and emerging trends. A dual-map overlay was constructed for journals. The parameters of CiteSpace were as follows: time slicing (2013 to 2022), years per slice (1), links (strength: cosine, scope: within slices), selection criteria (g-index: k=5 in country, institution, author collaboration network analysis and keyword co-occurrence), pruning (pathfinder, pruning sliced networks). Cluster labels used keywords by log likelihood ratio (LLR). To identify emerging topics, we detected keywords and references with a strong citation burst.

Ethical approval was waived by the local ethical board because the data came from public databases, and no human or animal subjects were involved.

## 3 Results

### 3.1 An overview of the annual growth trend

As shown in Figure 1A, from 2013 to 2022, a total of 8888 English-language articles met our inclusion criteria. The number of annual publications began to exceed 500 in 2016, and only after 3 years did the annual publications exceed 1000. The annual number of publications increased from 267 in 2013 to 1768 in 2022, with an amazing increase of over sixfold. We further analyzed the annual national output of the 10 most productive countries and regions (Figure 1B). The different colors represent different countries, and the slope represents the speed of annual publication number increase. China ranked first in the number of annual publications, followed by the United States and Spain. The top 10 publishers according to their contribution to the total number of articles about the effect of intestinal microbes on obesity are shown in Figure 1C. The different colors represent different years’ publications. Regarding the number of publications, Elsevier (1685) and Springer Nature (1388) far surpassed other publishers, confirming their unsurpassable international position in the publishing industry.

**FIGURE 1.**
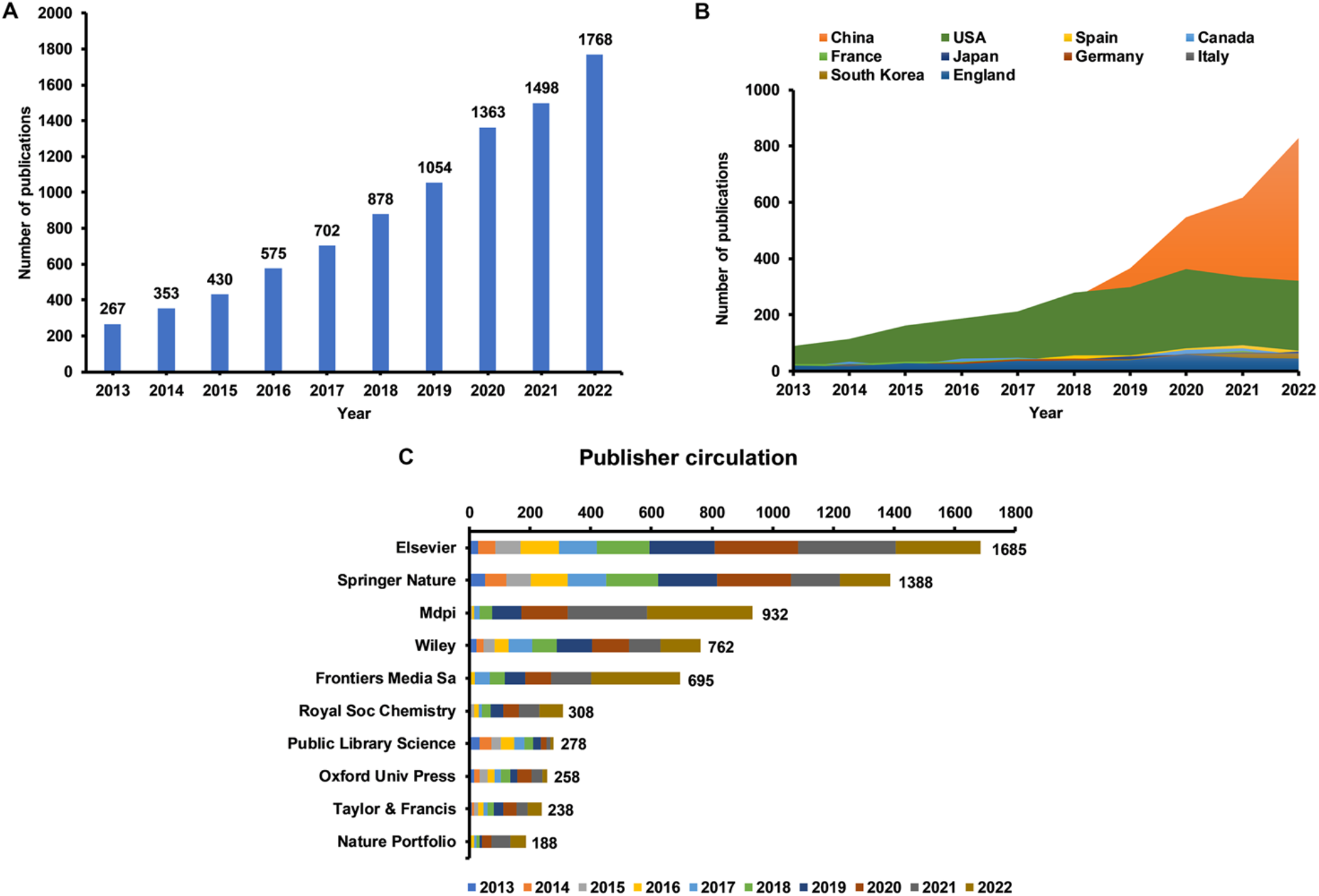
Global publication trend and country or region analysis for the effect of intestinal microbes on obesity research. (A) Annual worldwide publication output. (B) Growth trends in publication output from the top 10 most productive countries. (C) Top 10 publishers according to their contribution to the total number of articles on the effect of intestinal microbes on obesity research.

### 3.2 Country influence and collaboration

In the ten years from 2013-2022, 124 countries or regions participated in the domain of intestinal microbes and obesity. China ranked first with 3036 publications, accounting for 34.16% of all publications, followed by the United States (2356, 26.51%) and Spain (480, 5.40%) (Figure 2A, Table 1). Nevertheless, the United States tops the ranking of the most cited countries with a frequency (number of documents) of 87065, followed by China (65250), France (13809), and Canada (13496) (Supplementary Table S1). The country collaboration map shows the overall perspective of countries’ and regions’ academic cooperation (Supplementary Figure S1). The shade of blue represents the number of articles published, dark blue indicates more publications, and as the color becomes lighter, the publications decreased. The red line represents that there is academic collaboration between the linked two countries. The thicker the line is, the more cooperation two countries have. The United States had the broadest academic associations with other countries or regions, while its collaboration with China was the tightest, followed by Canada. The United States and China both had a frequent connection with European countries. Moreover, among European countries, there was also tight collaboration, but communication among the other countries or regions still needs to be strengthened.

**FIGURE 2.**
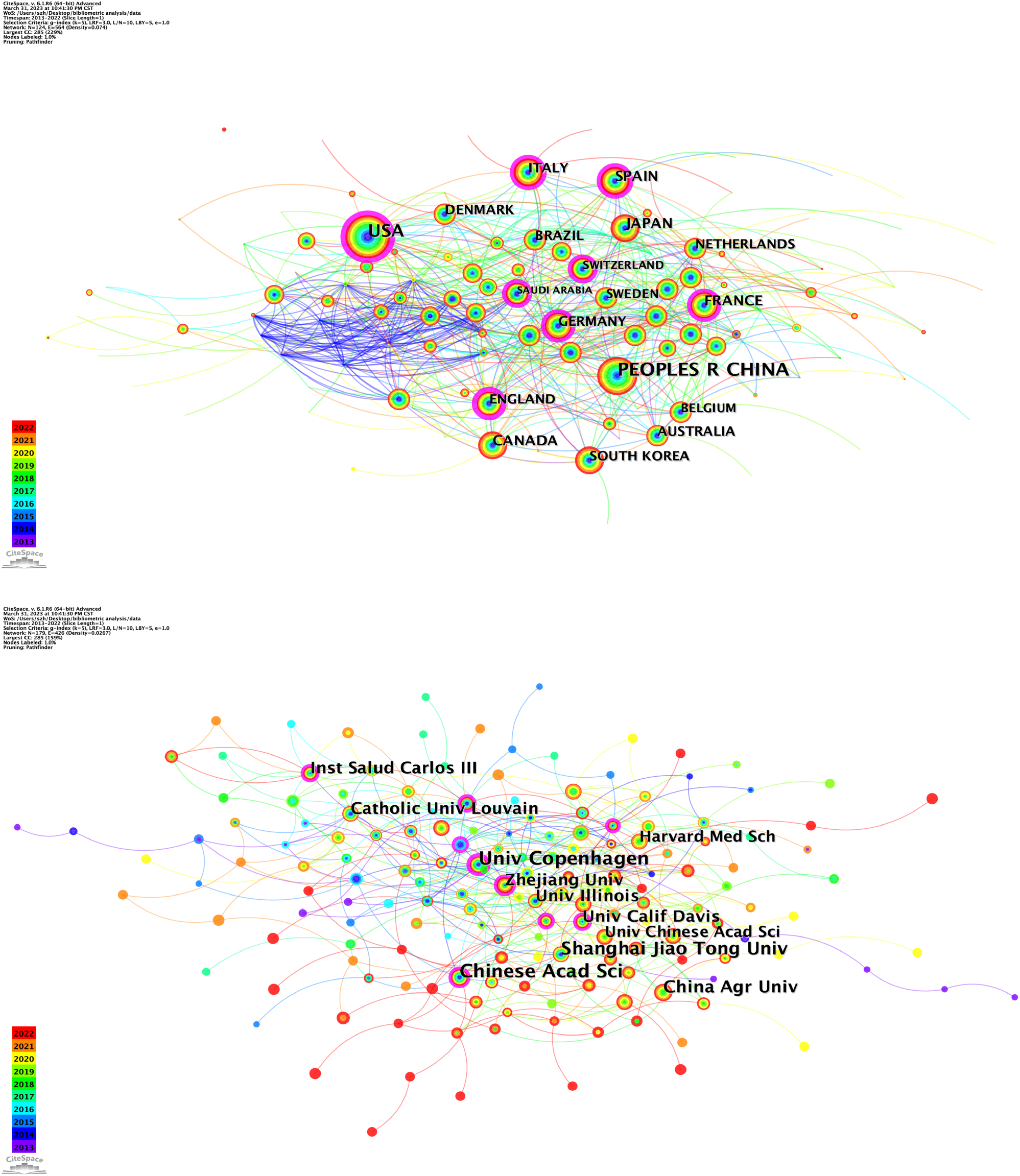
CiteSpace network visualization map of countries/regions and institutions. (A) CiteSpace network visualization map of countries/regions involved in the effect of intestinal microbes on obesity research (node label: by citation, label font size: proportional). (B) CiteSpace network visualization map of institutions involved in the effect of intestinal microbes on obesity research (node label: by citation, label font size: proportional).

**TABLE 1.** Top 10 productive countries/regions related to the effect of intestinal microbes on obesity.

| Rank | Countries | Articles | Centrality | Percentage(N/8888) |
| --- | --- | --- | --- | --- |
| 1 | China | 3036 | 0.02 | 34.16% |
| 2 | the United States | 2356 | 0.25 | 26.51% |
| 3 | Spain | 480 | 0.14 | 5.40% |
| 4 | Canada | 466 | 0.07 | 5.24% |
| 5 | France | 432 | 0.11 | 4.86% |
| 6 | Japan | 406 | 0.02 | 4.57% |
| 7 | Germany | 382 | 0.12 | 4.30% |
| 8 | Italy | 372 | 0.12 | 4.19% |
| 9 | South Korea | 364 | 0.05 | 4.10% |
| 10 | England | 355 | 0.23 | 3.99% |

There were 179 institutions included in the results of institution analysis. The top 10 institutions mostly came from China (5) and the USA (2), followed by Denmark (1), Belgium (1), and Spain (1). Chinese academic of sciences, the University of Copenhagen and Shanghai Jiao Tong University had the highest total link strength, revealing close cooperation with other institutions (Figure 2B, Table 2).

**TABLE 2.** Top 10 productive institutions related to the effect of intestinal microbes on obesity.

| Rank | Institutions | Articles | Centrality | Country |
| --- | --- | --- | --- | --- |
| 1 | University of Copenhagen | 209 | 0.28 | Denmark |
| 2 | Chinese academic of sciences | 202 | 0.18 | China |
| 3 | Shanghai Jiao Tong university | 174 | 0.09 | China |
| 4 | China Agricultural University | 129 | 0.04 | China |
| 5 | Catholic University of Louvain | 114 | 0.08 | Belgium |
| 6 | Zhejiang University | 107 | 0.14 | China |
| 7 | Inst Salud Carlos III | 94 | 0.10 | Spain |
| 8 | University of California, Davis | 93 | 0.11 | the United States |
| 9 | University of Illinois | 91 | 0.09 | the United States |
| 10 | University of Chinese academic of sciences | 90 | 0.02 | China |

### 3.3 Distribution and co-authorship of authors

In this analysis, 16 authors had published at least 21 publications. The 10 most productive authors are listed in Table 3. Most of the authors were from Europe, with Belgium (n=3), followed by China (n=3), Canada (n=2), Sweden (n=1), and France (n=1). Patrice D. Cani (97 publications) and Nathalie M. Delzenne (73 publications), both from Université catholique de Louvain, contributed the top two most publications, followed by Reimer, Raylene A (51 publications) from the University of Calgary. The top two authors maintained the most contact with other research labs, and the cooperation between Patrice D. Cani and Nathalie M. Delzenne was close (Figure 3A). Co-cited authors are shown in Figure 3B and Table 4. Peter J. Turnbaugh, and Patrice D. Cani rank the top two, which manifests their centrality in the research field.

**FIGURE 3.**
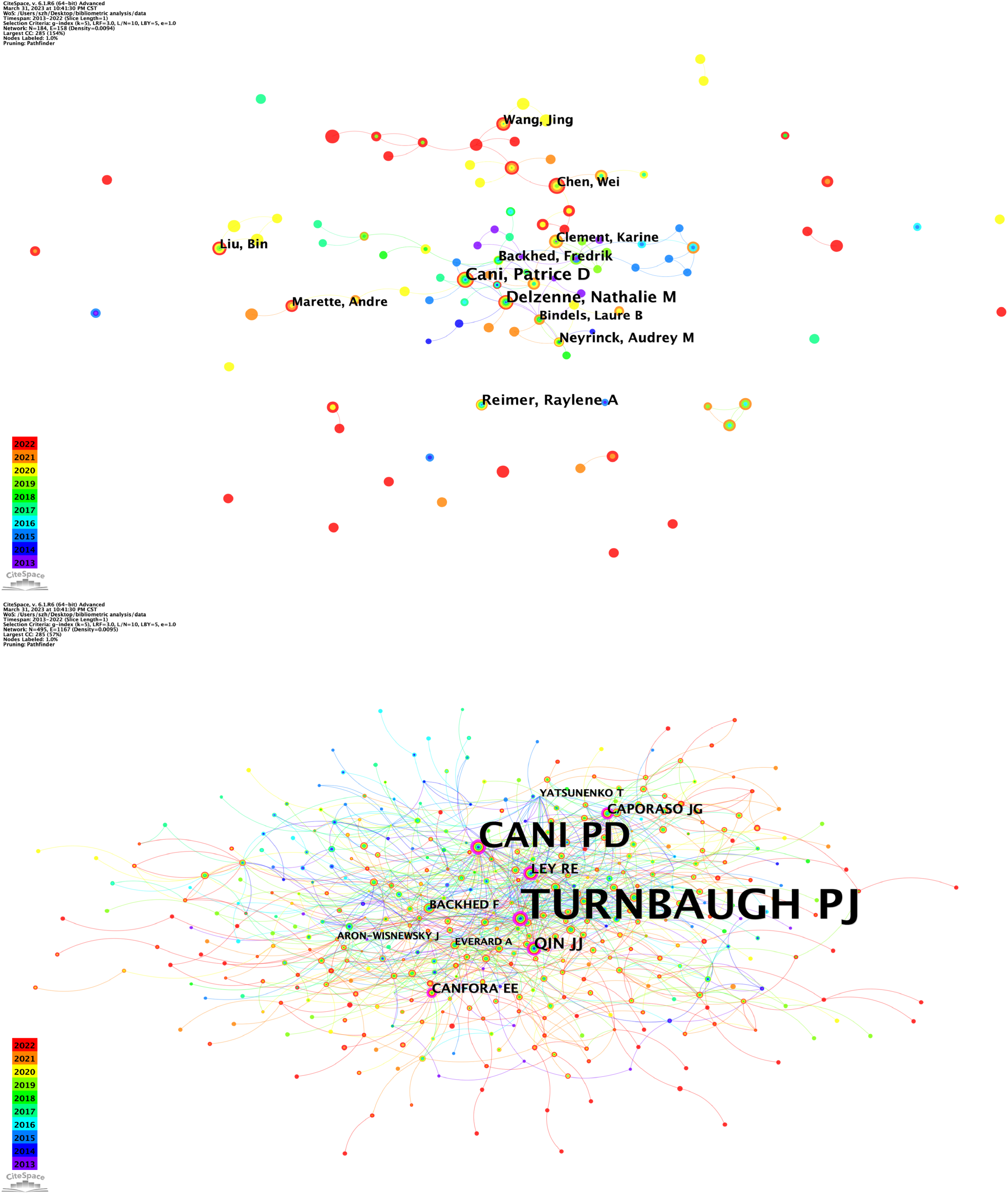
CiteSpace network visualization map of authors and co-cited authors. (A) CiteSpace network visualization map of authors involved in the effect of intestinal microbes on obesity research (node label: by citation, label font size: proportional). (B) CiteSpace network visualization map of co-cited authors involved in the effect of intestinal microbes on obesity research (node label: by centrality, label font size: proportional).

**TABLE 3.**
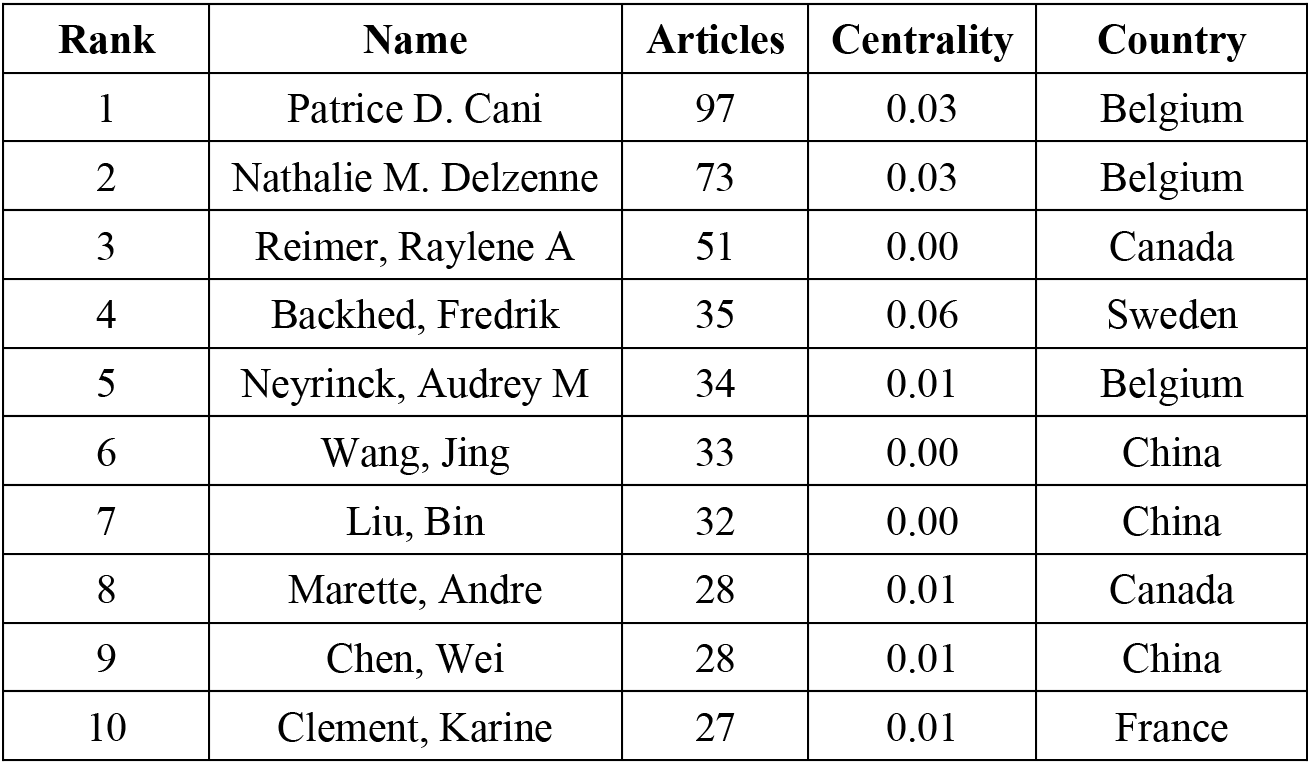
Top 10 productive authors related to the effect of intestinal microbes on obesity.

**TABLE 4.**
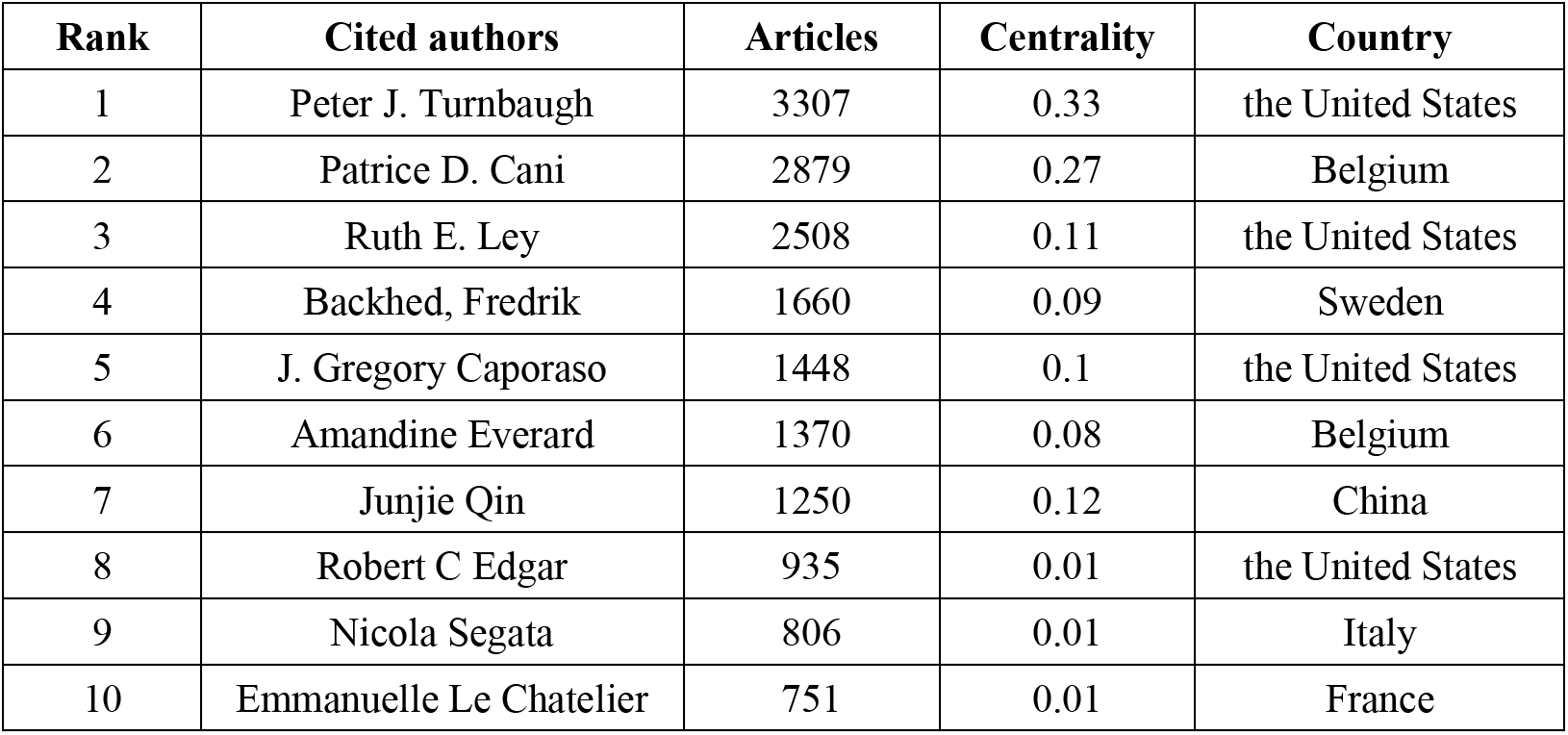
Top 10 co-cited authors related to the effect of intestinal microbes on obesity.

### 3.4 Analysis of journals and co-cited academic journals

As shown in Table 5, the top 10 most productive journals published 2155 publications, accounting for 24.25% of all publications. Nutrients (impact factor (IF) 2021, 6.706) published the most articles (399 publications, 4.49%), followed by Scientific Reports (IF 2021, 4.997; 339 publications, 3.81%) and Food & Function (IF 2021, 6.317; 281 publications, 3.16%). The top three highest co-cited journals were Nature (IF 2021, 69.504; 6446 citations), PLOS ONE (IF 2021, 3.752; 6247 citations), and Proceedings of the National Academy of Sciences of the United States of America (IF 2021, 12.779; 5329 citations). The co-citation relationship among different journals was visualized in a co-citation network (Figure 4, Table 6). Nature had the highest centrality (0.18), followed by PLOS ONE (0.17), which means that the published articles in the two journals were widely recognized and cited by researchers.

**FIGURE 4.**
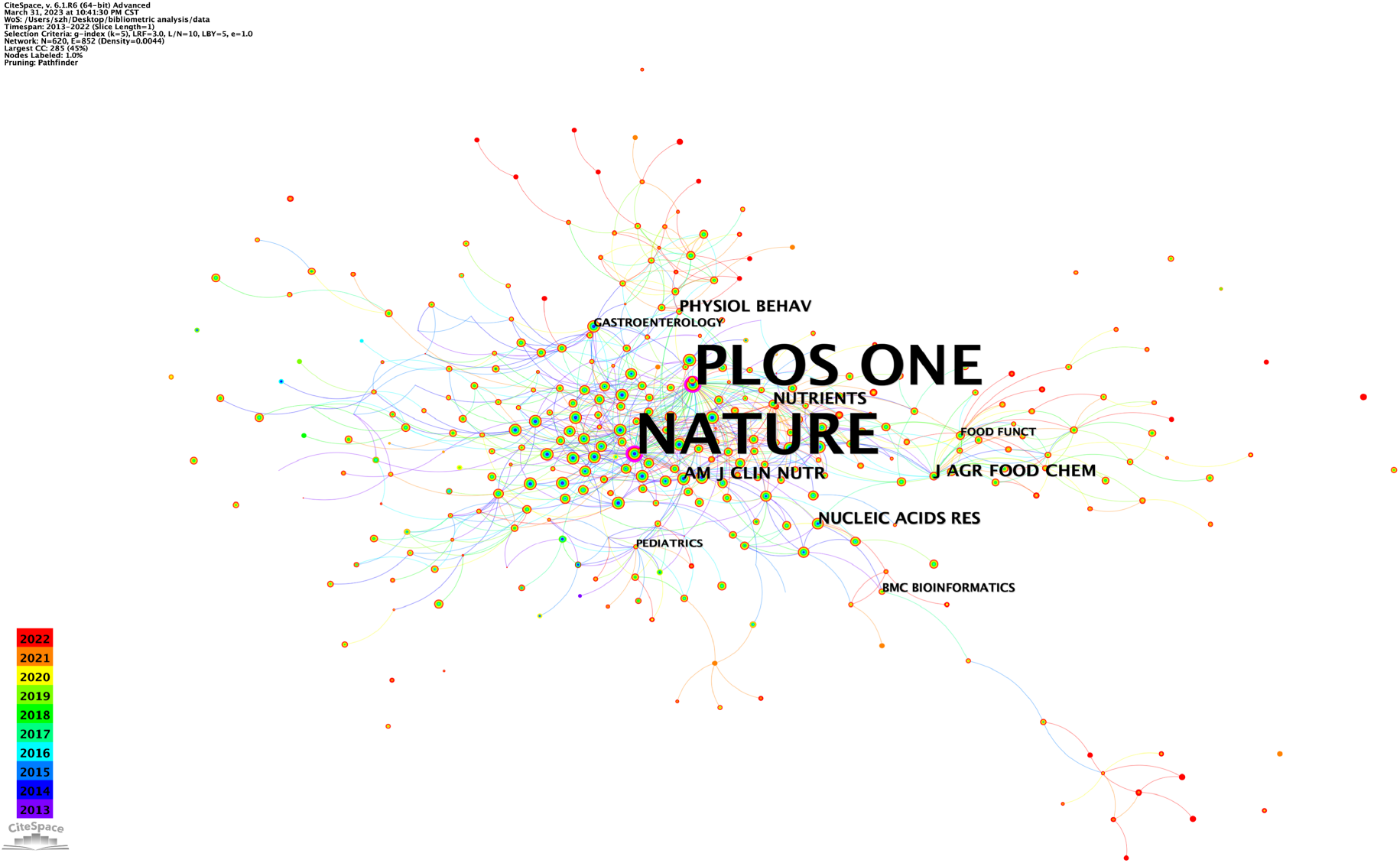
CiteSpace network visualization map of co-cited journals involved in the effect of intestinal microbes on obesity research (node label: by centrality, label font size: proportional).

**TABLE 5.** Top 10 productive journals related to the effect of intestinal microbes on obesity.

| Sources | Articles | Percentage(N/8888) | IF |
| --- | --- | --- | --- |
| Nutrients | 399 | 4.49% | 6.706 |
| Scientific Reports | 339 | 3.81% | 4.997 |
| Food & Function | 281 | 3.16% | 6.317 |
| PLOS ONE | 265 | 2.98% | 3.752 |
| Frontiers in microbiology | 226 | 2.54% | 6.064 |
| Journal of functional foods | 171 | 1.92% | 5.223 |
| Molecular nutrition & food research | 136 | 1.53% | 6.575 |
| Frontiers in nutrition | 116 | 1.31% | 6.590 |
| Journal of agricultural and food chemistry | 114 | 1.28% | 5.895 |
| International journal of molecular sciences | 108 | 1.22% | 6.208 |

**TABLE 6.**
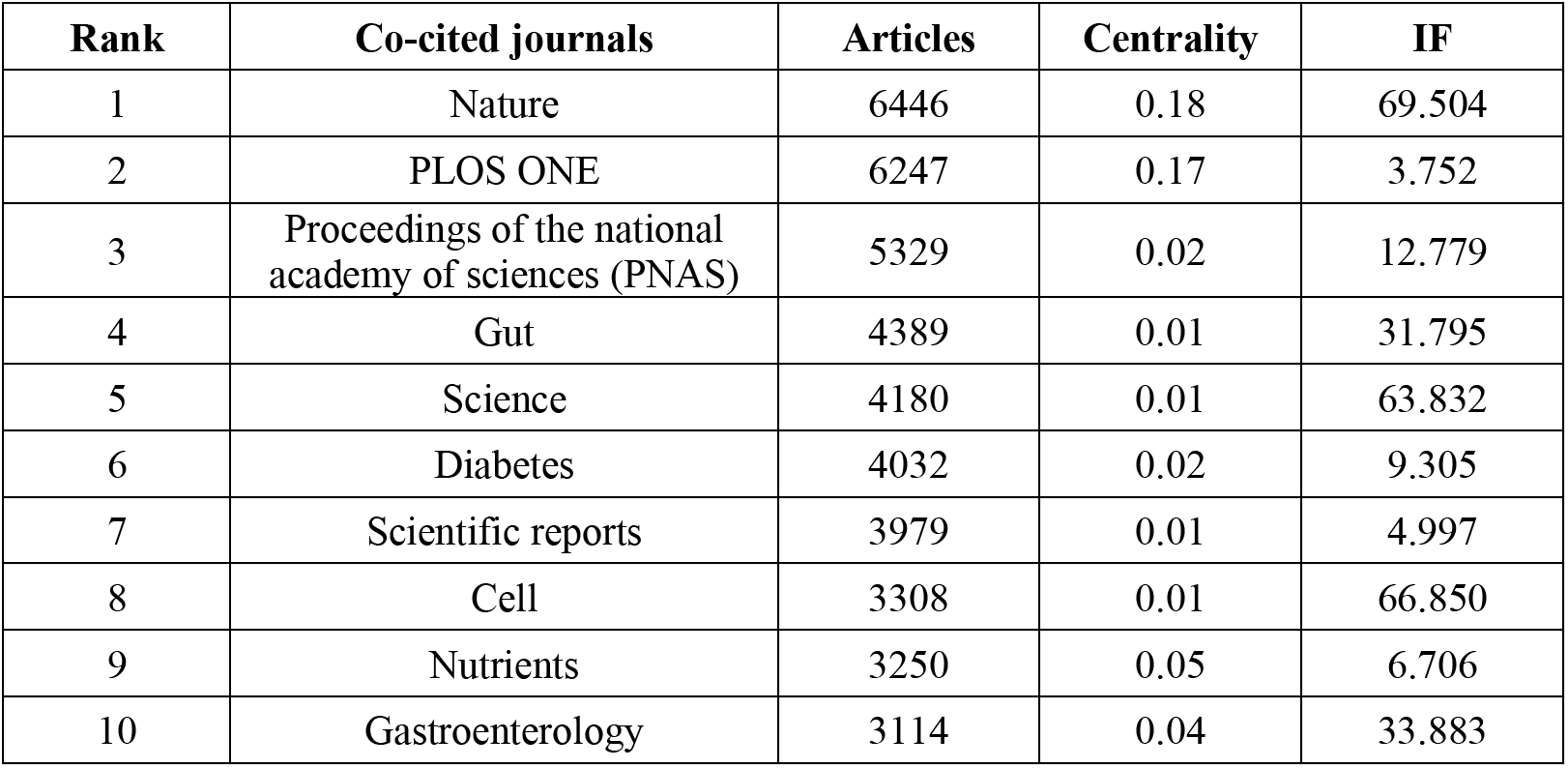
Top 10 co-cited journals related to the effect of intestinal microbes on obesity.

Supplementary Figure S2 shows the dual-map overlay of journals. The yellow, green, and orange spline waves represent citations made by the source articles. Each spline curve starts with the citing map on the left and points to the cited map on the right. The label represents the subject covered by the journal. The dual-map overlay shows seven main citation paths. The published articles were mostly focused on journals in the fields of medicine, medical, clinical, molecular, biology, and immunology and partly focused on veterinary, animal and science. Cited articles were mostly published in journals in the fields of molecular biology and genetics, and many articles were published in journals in the fields of health, nursing, medicine, environmental, toxicology and nutrition.

### 3.5 Analysis of keywords

#### 3.5.1 Co-occurrence analysis

Keywords could accurately reflect the main research point of an article, which has a high condensation in a research field and can directly point to the center of the text. Therefore, a high frequency of keywords represents hot issues in a research field and research hotspots. The keyword co-occurrence graph for the effects of intestinal microbes on obesity is shown in Figure 5. The density value was 0.0457. The ten most frequent keywords were gut microbiota, obesity, inflammation, insulin resistance, intestinal microbiota, health, metabolism, diet, gut microbiome, and metabolic syndrome (Table 7).

**FIGURE 5.**
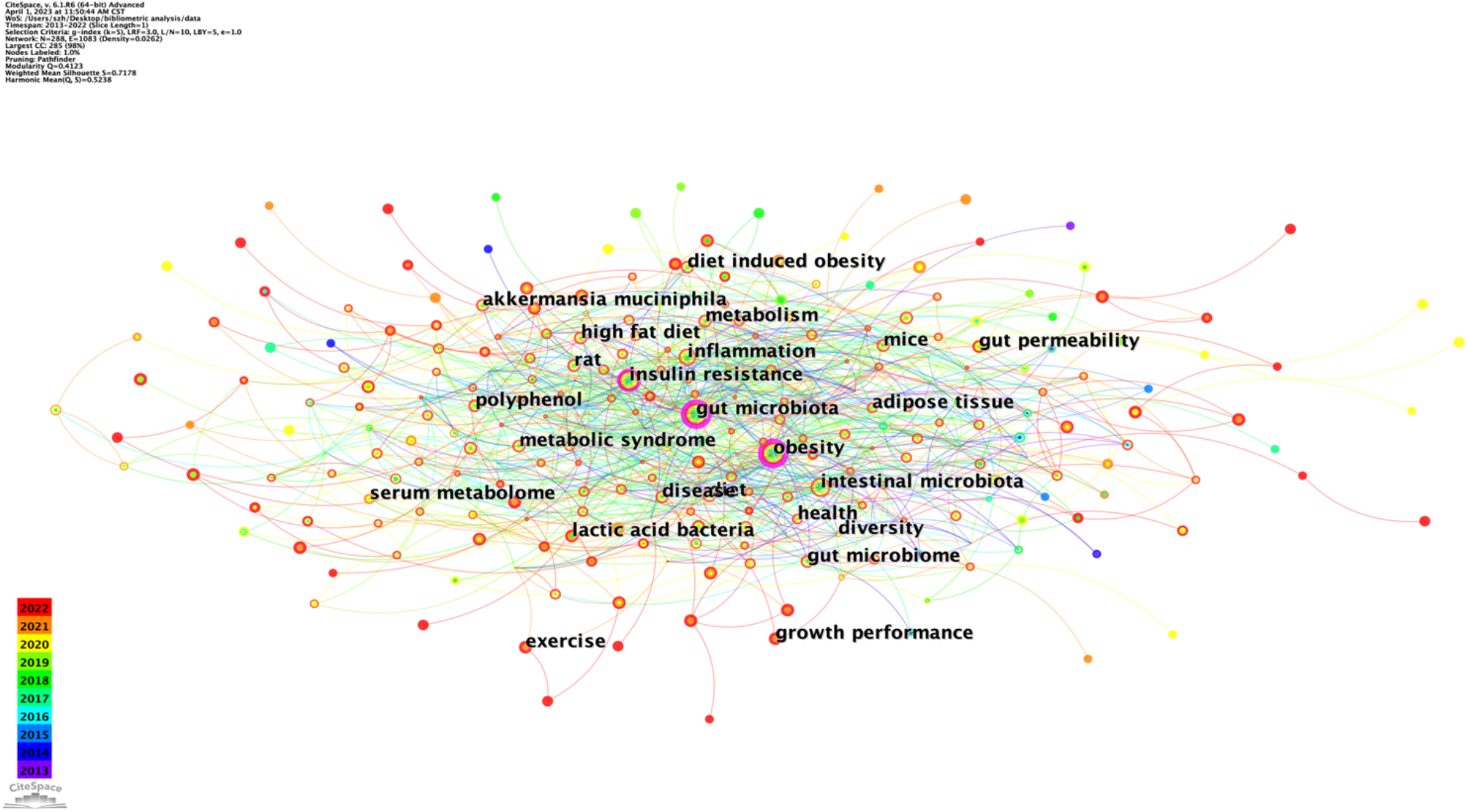
Keyword co-occurrence network for the effect of intestinal microbes on obesity research (node label: by centrality, label font size: uniformed).

**TABLE 7.** Top 10 keywords related to the effect of intestinal microbes on obesity.

| Rank | Keywords | Articles | Centrality |
| --- | --- | --- | --- |
| 1 | gut microbiota | 4673 | 0.39 |
| 2 | obesity | 3587 | 0.23 |
| 3 | inflammation | 1578 | 0.04 |
| 4 | insulin resistance | 1368 | 0.12 |
| 5 | intestinal microbiota | 1038 | 0.01 |
| 6 | health | 869 | 0.02 |
| 7 | metabolism | 862 | 0.02 |
| 8 | diet | 848 | 0.01 |
| 9 | gut microbiome | 825 | 0.01 |
| 10 | metabolic syndrome | 714 | 0.01 |

#### 3.5.2 Cluster analysis

The clustering analysis of the keywords yielded 288 nodes and 1083 links, with a Q value of 0.4123 (>0.3) and an S value of 0.7178 (>0.5), which are two important indicators to evaluate the significance of the clustering effect. There were 8 clusters, including diversity, insulin resistance, short-chain fatty acids, activation, food intake, bariatric surgery, body composition, and nonalcoholic fatty liver disease (Figure 6). “diversity” # 0 was the largest cluster, followed by “insulin resistance” # 1 and “short-chain fatty acids” # 2. Timeline View analysis was conducted to further analyze the keywords of the effects of intestinal microbes on obesity (Supplementary Figure S3).

**FIGURE 6.**
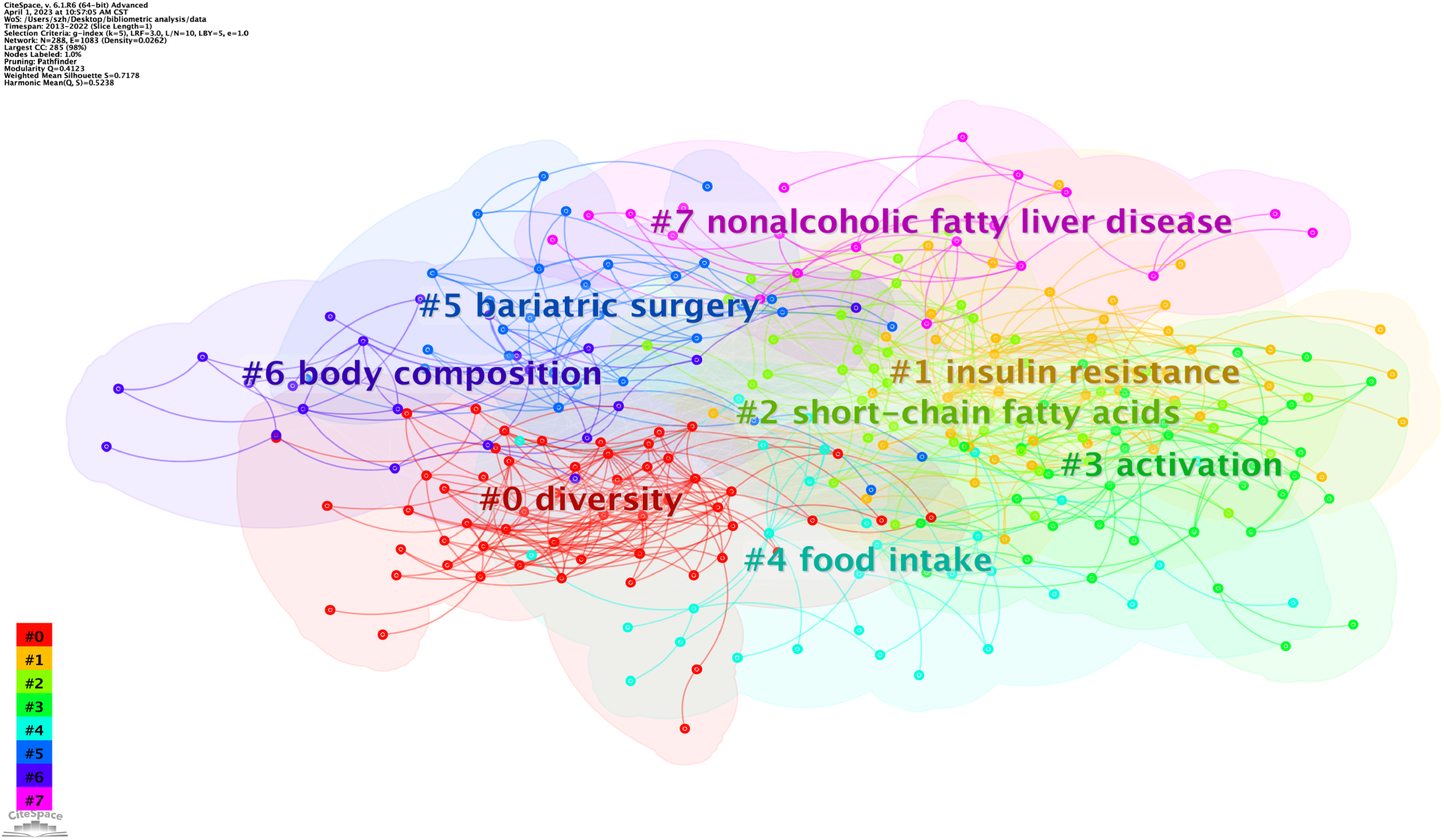
Keyword clusters for the effect of intestinal microbes on obesity research.

#### 3.5.3 Burst detection

CiteSpace was further used to perform burst detection with high frequency and therefore revealed research frontiers and hotspot trends over a period. As shown in Supplementary Table S2, the first 25 keywords were sorted by strengths of burst to discover the research hotspots about the effects of intestinal microbes on obesity. Keywords such as “ecology” (strength 28.33), “human gut microbiota” (strength 27.34), “microbiota” (strength 23.44), “diet induced obesity” (strength 21.12), and “irritable bowel disease” (strength 16) were the strongest keywords leading the research boom from 2013-2018. Keywords such as “obese” (strength 9.3) and “serum” (9.69) were the most recent trending keywords. Supplementary Table S3 shows the top 25 references with the strongest citation bursts, which were recognized as a critical milestone and led the development direction of the field for a while.

## 4 Discussion

In this study, we performed a bibliometric analysis of the effects of intestinal microbes on obesity-related studies from 2013 to 2022 using the core collection of WoSCC to comprehensively understand global research trends and hotspots and provide references for researchers in this field or those who want to become involved in the field. Based on the above analysis, it is certain that obesity is closely influenced by intestinal microbes.

### 4.1 Global trends in effects of intestinal microbes on obesity

The amount of annual scientific production is an important indicator of development in an academic field. In our study, a total of 8888 original articles from 1392 journals met the inclusion criteria. The article annual growth rate reached 22.93%, and the annual growth rate from 2019 to 2020 was the highest. An article published by Lawrence A. David et al., which was the most co-cited, found that an altered diet could lead to a rapid gut microbiome response. The results showed that short-term consumption of diets composed entirely of animal or plant products alters microbial community structure and overwhelms interindividual differences in microbial gene expression (DavidMaurice et al., 2014). The second most cited article was published by Amandine Everard et al. They isolated a mucin-degrading bacterium that resides in the mucus layer, named *Akkermansia muciniphila*. The results showed that the abundance of *A. muciniphila* decreased in obese and type 2 diabetic mice, but prebiotic feeding could normalize *A. muciniphila* abundance. Moreover, high-fat diet-induced metabolic disorders, including fat-mass gain, metabolic endotoxemia, adipose tissue inflammation, and insulin resistance, could be reversed by *A. muciniphila* treatment (EverardBelzer et al., 2013). Although these two articles are about different research, they both proved the correlations between intestinal microbes and the host, which have greatly driven the development of the field.

Among the 10 most influential journals in effects of intestinal microbes on obesity research, four were also among the top 10 co-cited journals (Nature, Proceedings of the national academy of sciences (PNAS), Gut, Science). Among these, the IF value of Nature was 69.504 (2021), followed by Science (63.832), Gut (31.795) and PNAS (12.779), indicating their outstanding contributions in the field of effects of intestinal microbes on obesity research. The most frequently co-cited journals were Nature, PLOS ONE, PNAS, and Gut. Notably, three of the top 10 co-cited references were published in Nature, one of them was published in PNAS, and one of them was published in Gut. The main research hotspots were about the mechanism of dietary intervention or human genetics to intestinal microbes. Moreover, richness or kinds of intestinal microbes can also modulate the host’s metabolism, such as obesity or type 2 diabetes. These findings have laid the foundation for a deeper understanding of the regulatory relationship between intestinal microbes and obesity or type 2 diabetes.

### 4.2 Co-authorship networks in effects of intestinal microbes on obesity research

The closeness of collaboration between countries/regions, institutions, and authors was assessed, which can help find the laws of scientific research cooperation, guiding more effective scientific research activities, and promoting potential collaborative opportunities for other groups. According to Table 1, China, with 3036 articles, ranks first among the top 10 total publication countries, followed by the United States, with 2356 publications. Notably, the total number of publications in the USA (87065) was greater than that in China (65250) and far greater than that in France (13809), Canada (13496) and the other top 10 most cited countries. The total citations of the United States and China are far more than several other top 10 most cited countries citations combined. Without doubt, the United States and China demonstrate their position as leaders in the field. Notably, Israel with 49 publications was the most cited country per article (average article citations, 124.18), suggesting that the quality of research on effects of intestinal microbes on obesity in Israel is very high, the similar story is unfolding in many European countries such as Belgium (average article citations, 107.85), Sweden (average article citations, 103.95), etc., which sets a model for other countries/regions in this research field (Supplementary Table S1, S4). To improve the quality of articles, the reliability of the data source and the rigor of the experimental design may be the first factors affecting the quality of publications, and more attention should be given to these aspects in future studies. According to Figure 2B, the bottom right is most dominated by universities from China, while the upper left is most dominated by schools in North America or Europe. Nevertheless, more cooperation between the two clusters could be conducted to catalyze breakthrough progress in research on the effects of intestinal microbes on obesity.

Four of the top 10 institutions are based in China, followed by the United States with three, which are thereby maximizing regional advantages and demonstrating the dominance of the United States and China in the field. This may partly explain why China and the United States consistently maintain a high quantity of publications. University of Copenhagen in Denmark was the most productive institution worldwide, followed by Chinese academic of sciences in China, indicating that these two institutions participated in the most collaborations with other institutions worldwide. Although Spain and Canada ranked third and fourth in terms of total publications, only one of the Spanish research institutions ranked in the top 10, indicating a lack of institutions with professional and research stature in terms of the effects of intestinal microbes on obesity research (Table 1-2). The most effective organizations and groups are leading the trends in the effects of intestinal microbes on obesity research; thus, further study at these institutions will ensure continuous future development in this field.

### 4.3 Basic knowledge and hotspots in effects of intestinal microbes on obesity research

#### 4.3.1 Basic knowledge

The most frequently cited references possess pivotal academic position in the field. Either positive or negative conclusions from the documents influence others’ research direction. In our study, the top co-cited references were used to investigate the knowledge base for the effects of intestinal microbes on obesity research. Among the top 10 co-cited references, we found that second-generation sequencing was a vital technology that relies on its high-throughput characteristic, making it easy to sequence the transcriptome or genome of a species (Schuster, 2008). With the development of sequencing technology, a large amount of data is generated. To make more accurate and full use of sequencing data, software and data analysis platforms have also been developed. An open-source software package named DADA2 is used for modeling and correcting Illumina-sequenced amplicon errors. With the help of this software, researchers could accurately reconstruct amplicon-sequenced communities at the highest resolution, which ensured the accuracy of the research to the greatest extent (CallahanMcMurdie et al., 2016). Another utility tool named QIIME 2 could serve not only as a marker-gene analysis tool but also as a multidimensional and powerful data science platform that can be rapidly adapted to analyze diverse microbiome features (BolyenRideout et al., 2019). These tools help drive rapid development in microbiome research. With the help of sequencing, recent research found that the richness of the human gut microbiome correlated with human metabolic markers. Individuals with a low bacterial richness were characterized by more marked overall adiposity, insulin resistance and dyslipidemia and a more pronounced inflammatory phenotype when compared with high bacterial richness individuals (Le ChatelierNielsen et al., 2013). Correspondingly, with higher gut microbiome gene richness and *A. muciniphila* abundance could exhibit the healthiest metabolic status, particularly in fasting plasma glucose, plasma triglycerides and body fat distribution (DaoEverard et al., 2016). The obesity-associated gut microbial species *Bacteroides thetaiotaomicron,* a glutamate-fermenting commensal, is linked to changes in circulating metabolites, and the abundance of *Bacteroides thetaiotaomicron* is markedly decreased in obese individuals and inversely correlated with serum glutamate concentration (LiuHong et al., 2017). Human obesity is a heterogeneous condition in the context of pathogenesis, pathophysiology and therapeutic responsiveness. Studies of alterations in the genome—the microbial gut metagenome—may define subsets of adult individuals with different metabolic risk profiles, which could contribute to resolving some of the heterogeneity associated with adiposity-related phenotypes (Le ChatelierNielsen et al., 2013). All these findings suggest that gut microbiome richness is a key factor in maintaining the homeostasis of human health.

The gut microbiome can rapidly respond to an altered diet, and a dynamic balance can be achieved between intestinal microbes and diet immediately, indicating that not only a long-term diet but also a short-term diet can influence the gut microbiome (DavidMaurice et al., 2014). Weight-loss intervention by bariatric surgery partially reversed obesity-associated microbial and metabolic alterations in obese individuals (LiuHong et al., 2017). This means that external interventions can affect intestinal microbes. Research also proved that transmissible and modifiable interactions between diet and microbiota influence host biology. The transformation correlated with invasion of members of *Bacteroidales* from Ln into Ob microbiota that prevented development of increased adiposity and body mass in Ob cage mates and transformed their microbiota’s metabolic profile to a leanlike state. Therefore, it may be possible to intervene in obesity by targeting the gut microbiota (RidauraFaith et al., 2013), and there is already evidence proving this speculation. *Akkermansia muciniphila* is a mucin-degrading bacterium that resides in the mucus layer. Research found that *A. muciniphila* decreased in obese and type 2 diabetic mice, and *A. muciniphila* treatment reversed high-fat diet-induced metabolic disorders, including fat-mass gain, metabolic endotoxemia, adipose tissue inflammation, and insulin resistance (EverardBelzer et al., 2013). Pasteurized *A. muciniphila* could enhance its capacity to reduce fat mass development, insulin resistance and dyslipidemia. Moreover, Amuc_1100, a purified membrane protein from *Akkermansia muciniphila*, could improve the gut barrier and partly recapitulate the beneficial effects of the bacterium (PlovierEverard et al., 2017). All these findings are of great driving significance, indicating that it is feasible to use microbes to treat obesity, but more research is needed to support this hypothesis.

In addition to the underlying effect of intestinal microbes on obesity, substantial evidence has suggested that intestinal microbes also have an effect on type 2 diabetes, which is also a major public health issue throughout the world. A metagenome-wide association study (MGWAS) was used to identify and validate type-2-diabetes-associated markers and to establish the concept of a metagenomic linkage group, which enabled taxonomic species-level analyses. Type 2 diabetes is accompanied by a decrease in the abundance of some universal butyrate-producing bacteria and an increase in various opportunistic pathogens (QinLi et al., 2012). Conversely, high bacterial richness was related to a reduction in insulin resistance (Le ChatelierNielsen et al., 2013), and *A. muciniphila* treatment could also reverse insulin resistance (EverardBelzer et al., 2013) (Table 8).

**TABLE 8.** The top 10 co-cited references related to the effect of intestinal microbes on obesity.

| Rank | Title | Author | Article type | Journal | DOI | Articles |
| --- | --- | --- | --- | --- | --- | --- |
| 1 | Diet rapidly and reproducibly alters the human gut microbiome | Lawrence A. David et al., 2013 | Letter | Nature | 10.1038/nature12820 | 383 |
| 2 | Cross-talk between *Akkermansia muciniphila* and intestinal epithelium controls diet-induced obesity | Amandine Everard et al., 2013 | Article | PNAS | 10.1073/pnas.1219451110 | 367 |
| 3 | *Akkermansia muciniphila* and improved metabolic health during a dietary intervention in obesity-relationship with gut microbiome richness and ecology | Maria Carlota Dao et al., 2016 | Article | Gut | 10.1136/gutjnl-2014-308778 | 303 |
| 4 | A metagenome-wide association study of gut microbiota in type 2 diabetes | Junjie Qin et al., 2012 | Article | Nature | 10.1038/nature11450 | 291 |
| 5 | Richness of human gut microbiome correlates with metabolic markers | Emmanuelle Le Chatelier et al., 2013 | Article | Nature | 10.1038/nature12506 | 285 |
| 6 | Gut microbiota from twins discordant for obesity modulate metabolism in mice | Vanessa K. Ridaura et al., 2013 | Article | Science | 10.1126/science.1241214 | 276 |
| 7 | A purified membrane protein from *Akkermansia muciniphila* or the pasteurized bacterium improves metabolism in obese and diabetic mice | Hubert Plovier et al., 2017 | Letters | Nature Medicine | 10.1038/nm.4236 | 275 |
| 8 | DADA2: High-resolution sample inference from Illumina amplicon data | Benjamin J Callahan et al., 2016 | Brief communications | Nature methods | 10.1038/nmeth.3869 | 273 |
| 9 | Reproducible, interactive, scalable and extensible microbiome data science using QIIME 2 | Evan Bolyen et al., 2019 | Correspondence | Nature biotechnology | 10.1038/s41587-019-0209-9 | 265 |
| 10 | Gut microbiome and serum metabolome alterations in obesity and after weight-loss intervention | Ruixin Liu et al., 2017 | Article | Nature medicine | 10.1038/nm.4358 | 264 |

#### 4.3.2 Research hotspots

Keywords with high frequency are usually used to accurately reveal the main research interests and hotspots in the field within a certain period, which help researchers to precisely catch up to the research trends from numerous studies. By analyzing the co-occurrence keywords, “gut microbiota” and “obesity” had the highest frequency and centrality ranking the top two, indicating that the main research interests focused on the relationship between “gut microbiota” and “obesity”. The effects of intestinal microbes on obesity have been supported by several trials, which have all shown that intestinal microbes modulate the metabolism of obesity (EverardBelzer et al., 2013; Le ChatelierNielsen et al., 2013; RidauraFaith et al., 2013). The clustering function was used to show the whole network map. The 8 clusters of keywords presented in this paper illustrate 8 research topics, which to some extent represent 8 significant aspects of the research. Studies in the same cluster might have similar research themes. As shown in Figure 6, the top two largest clusters, “diversity” # 0 and “insulin resistance” # 1, showed that the two topics were still the main themes in the effect of intestinal microbes on obesity research. Simultaneously, a timeline view analysis was conducted to further observe the evolution pace of each cluster and recognize the pivotal research direction through a microperspective. We can clearly observe in Supplementary Figure S3 that the research focus has shifted with time; by 2020, the research on several topics was more balanced. After 2020, the research in the “shot-chain fatty acids” #2 and “activation” #3 directions gradually weakened. The results indicated that gut microbiome research and the treatment of obesity and obesity-related diseases have become the main themes.

Furthermore, burst detection was an analytic tool that could effectively capture the dramatic increases in references or keywords in one research field within a specified period; therefore, it served as an important indicator of research hotspots or research frontiers over time. The evolution of the burst keywords over the last 10 years is shown in Supplementary Table S2. According to the ranking by burstiness strength, we observed that the top 15 keywords started to burst in 2013, and “ecology” remained the longest with an end time in 2018. More importantly, the “obese patient” and “serum” burst separately in 2019 and 2020 and continue to burst until now, which indicated that the two topics have received continuous attention in recent years and might be the main trends of research on the effect of intestinal microbes on obesity. Notably, there was a sudden and rapid growth in the number of articles in the field, which might be related to the term “serum”, which was only a new burst in that year.

Bariatric surgery is considered the only effective and sustainable weight loss method for obese patients. There is a 50–70% decrease in body weight and fat mass after surgical procedures such as Roux-Y gastric bypass (RYGB) and sleeve gastrectomy (SG) (PaganelliLuyer et al., 2019). Nevertheless, obese patients undergoing bariatric surgery may experience overgrowth of small intestinal bacteria (SabateCoupaye et al., 2017), a condition that snags with weight loss and increases the risk of micronutrient deficiency that appears to be harmful for the configuration and composition of intestinal microbiota (HibberdWu et al., 2017; Mach and Clark, 2017). To maintain the effect of surgery and avoid weight rebound, it is important for obese patients to correct the microbial balance and improve microbiota-host interactions with specific interventions (Aron-WisnewskyPrifti et al., 2019). Research has also shown that the elevated pH resulting from RYGB could ensure the survival of probiotic bacteria, making it possible for surgical patients to receive probiotic therapy (80). Probiotics are a kind of active microorganism beneficial to the host that colonizes the human intestinal tract and reproductive system, and they can improve the host microecological balance and play a beneficial role. Bariatric surgery is often followed by an increase in *Streptococcaceae* and a decline in *Bifidobacteriaceae* (PaganelliLuyer et al., 2019). Proper supplementation with probiotics can compensate for the intestinal microbial imbalance caused by surgery. Moreover, recent research showed that probiotic intervention could increase the levels of peptide YY and GLP-1 in mice (TianLi et al., 2016), reduce the level of intestinal inflammation (TsaiLin et al., 2019), and induce the production of anti-inflammatory cytokines (FolignéParayre et al., 2016). Meanwhile, intestinal peptide signals activate the gut-brain axis, which, in turn, exerts endocrine effects on other organ systems, especially the brain, regulating appetite, metabolism, and other dietary behaviors(Bliss and Whiteside, 2018; Hussain and Bloom, 2013; NeyrinckVan Hée et al., 2012; RastelliVan Hul et al., 2020). Consequently, weight loss and reduced insulin resistance occur. However, due to the heterogeneity of experimental techniques, methods and objects, the effectiveness and safety of microbial product intervention in improving obesity against intestinal microbiota need to be further verified, and the mechanism is not clear. Few studies have focused on the development of new functional microbiological products, and most of them have been carried out in mice. Before microbial products are reasonably and effectively treated for obesity, a large number of studies are needed, especially randomized controlled clinical trials.

Serum samples are a less invasive biomatrix that reflects the dynamic changes in the metabolome of the whole organism (DunnBroadhurst et al., 2011). The serum metabolome has emerged as a technique that focuses on defining the functional status of host-microbial relationships in biological specimens, which can reflect the dynamic changes in metabolites and explore disease-related metabolites or dysregulated metabolic pathways (metabolome analysis for investigating host-gut microbiota interactions). A top 10 co-cited reference showed the gut microbiome and serum metabolome alterations in obesity that patients after bariatric surgery not only appeared partially reversed obesity-associated microbial but also accompanied by metabolic alterations, including the decreased abundance of *B. thetaiotaomicron* and the elevated serum glutamate concentration, suggesting that it may be possible to intervene in obesity by targeting the gut microbiota (LiuHong et al., 2017). There are complex interactions between the host and intestinal microbes in carbohydrate, amino acid, lipid and nucleic acid metabolism. Intestinal microbes can use their respective metabolites to maintain intestinal viability while affecting the development, homeostasis and function of the host immune system through nutrition- and metabolite-dependent mechanisms (Brestoff and Artis, 2013). Small molecules, such as vitamins, fatty acids, amino acids, and bile acids, regulate host-intestinal metabolic homeostasis by binding to specific host membranes or nuclear receptors (HustedTrauelsen et al., 2017; PutignaniDel Chierico et al., 2016). The references with the highest citation bursts are unique contributions to the field of study. We found that articles related to bile acids have become the most cited articles in recent years (KohDe Vadder et al., 2016; WangLiao et al., 2019). Bile acids directly and rapidly affect the metabolism of bacteria, including membrane damage and disruption of amino acid, nucleotide and carbohydrate metabolism, and short-term exposure to bile acids significantly affects host metabolism by altering the bacterial community structure (TianGui et al., 2020). Another study showed that the gut microbiota also produced many other metabolites with important functions, such as short-chain fatty acids (SCFAs) and bile acids (KrishnanDing et al., 2018). Short-chain fatty acids can directly activate G-coupled receptors, inhibit histone deacetylases, and serve as energy substrates (KohDe Vadder et al., 2016). They thus affect various physiological processes and may contribute to health and disease. According to the above analysis, although notable advances in the effects of intestinal microbes on obesity have been made, our understanding of the interrelationships between them remains descriptive, and we still have numerous gaps to fill.

### 4.4 Strengths and Limitations

The bibliometric study conducted a systematic analysis of the basic situation, research hotspots, and trends in effects of intestinal microbes on obesity from a visualization perspective. The results of the bibliometric study were objective and accurate, which could provide a comprehensive guide for academics who are already or wish to work in this field. Nevertheless, there are still some limitations in our study. First, owing to the nature of the CiteSpace software, our data are filtered only from the WoSCC database, which is not sufficiently comprehensive and may lead to data omission. Second, our results were processed by CiteSpace software with certain algorithms, which could lead to bias in some of the results. Finally, only English articles were included from the database and analysis, potentially leading to a source bias.

## 5 Conclusion

Intestinal microbes possess essential research value and application prospects in obesity therapy. Compared with other review articles, the contribution of the study is evident in its visualized ways to reveal the effect of intestinal microbes on obesity. Research on the effects of intestinal microbes on obesity is increasing rapidly, highlighted by the outstanding contributions of China, the United States and Spain to the development of this field. Current research hotspots focus on related mechanisms of the effects of intestinal microbes on obesity and therapeutic methods for obesity with intestinal microbes. Diet, probiotic preparations and regulation of the intestinal flora, fecal microbe transplantation may have significant impacts on reducing the obesity epidemic. “Obese patient” and “serum” have become the latest burst that have attracted attention in recent years and will serve as the focus of future research. More cooperation and communication between countries and institutions should be strengthened to promote development in this field and benefit more patients with obesity.Data availability statement

The original contributions presented in the study are included in the article, and further inquiries can be directed to the corresponding author.

## Author Contributions

ZS and JG conceived the study. XY guided the methodological process. ZS, CT participated in data collection and analysis. ZS wrote the manuscript. GW, JG and XY revised the manuscript. All authors contributed to the article and approved the submitted version.

## Data Availability

All data produced in the present study are available upon reasonable request to the authors.

## Acknowledgments

The authors are indebted to Prof. Zhiyong Zong, who contributed his time, knowledge, and energy to developing this document.

## Publisher’s note

All claims expressed in this article are solely those of the authors and do not necessarily represent those of their affiliated organizations or those of the publisher, the editors and the reviewers. Any product that may be evaluated in this article or claim that may be made by its manufacturer is not guaranteed or endorsed by the publisher.

## Supplementary Material

The Supplementary Material for this article can be found online at:

