## Supplementary Table S1-S4, Supplementary Table Figure S1-S3 for "Study of the Effect of Intestinal Microbes on Obesity: A Bibliometric Analysis"

Supplementary Material

SUPPLEMENTARY TABLE S1 Top 10 most local cited countries/regions related to the effect of intestinal microbes on obesity.

| **Country** | **Total citations** | **Citations per article** |
| --- | --- | --- |
| the United States | 87065 | 52.20 |
| China | 65250 | 22.42 |
| France | 13809 | 52.71 |
| Canada | 13496 | 43.68 |
| Spain | 12780 | 35.01 |
| Belgium | 11648 | 107.85 |
| Japan | 10641 | 32.25 |
| Germany | 10155 | 59.04 |
| Korea | 9568 | 28.99 |
| Netherlands | 9542 | 53.61 |

SUPPLEMENTARY TABLE S2 Top 25 burst keywords in articles related to the effect of intestinal microbes on obesity. Burst strength indicates the importance of keywords to the research field. The years between “Begin” and “End” represent the period when the keyword was more influential. The thick dark bar represents the year in which the burst keyword appeared and the burst duration.

| **Keywords** | **Year** | **Strength** | **Begin** | **End** | **2013 - 2022** |
| --- | --- | --- | --- | --- | --- |
| ecology | 2013 | 28.33 | 2013 | 2018 | ▃▃▃▃▃▃▂▂▂▂ |
| human gut microbiota | 2013 | 27.34 | 2013 | 2017 | ▃▃▃▃▃▂▂▂▂▂ |
| microflora | 2013 | 23.44 | 2013 | 2016 | ▃▃▃▃▂▂▂▂▂▂ |
| diet induced obesity | 2013 | 21.12 | 2013 | 2014 | ▃▃▂▂▂▂▂▂▂▂ |
| inflammatory bowel disease | 2013 | 15.88 | 2013 | 2018 | ▃▃▃▃▃▃▂▂▂▂ |
| diversity | 2013 | 13.38 | 2013 | 2015 | ▃▃▃▂▂▂▂▂▂▂ |
| weight lo | 2013 | 12.42 | 2013 | 2014 | ▃▃▂▂▂▂▂▂▂▂ |
| escherichia coli | 2013 | 11.85 | 2013 | 2017 | ▃▃▃▃▃▂▂▂▂▂ |
| crohns disease | 2013 | 11.39 | 2013 | 2016 | ▃▃▃▃▂▂▂▂▂▂ |
| fecal microbiota | 2013 | 11.39 | 2013 | 2017 | ▃▃▃▃▃▂▂▂▂▂ |
| gene expression | 2013 | 11.28 | 2013 | 2016 | ▃▃▃▃▂▂▂▂▂▂ |
| endotoxemia | 2013 | 10.82 | 2013 | 2016 | ▃▃▃▃▂▂▂▂▂▂ |
| glucagon like peptide 1 | 2013 | 10.53 | 2013 | 2015 | ▃▃▃▂▂▂▂▂▂▂ |
| bacteria | 2013 | 10.14 | 2013 | 2015 | ▃▃▃▂▂▂▂▂▂▂ |
| ulcerative coliti | 2013 | 9.70 | 2013 | 2016 | ▃▃▃▃▂▂▂▂▂▂ |
| flora | 2013 | 9.55 | 2013 | 2014 | ▃▃▂▂▂▂▂▂▂▂ |
| intestinal permeability | 2014 | 12.25 | 2014 | 2016 | ▂▃▃▃▂▂▂▂▂▂ |
| gastrointestinal tract | 2014 | 10.65 | 2014 | 2017 | ▂▃▃▃▃▂▂▂▂▂ |
| intestinal microbiota | 2013 | 10.04 | 2014 | 2015 | ▂▃▃▂▂▂▂▂▂▂ |
| irritable bowel syndrome | 2015 | 16.00 | 2015 | 2018 | ▂▂▃▃▃▃▂▂▂▂ |
| human gut microbiome | 2016 | 12.08 | 2016 | 2018 | ▂▂▂▃▃▃▂▂▂▂ |
| immune system | 2017 | 12.76 | 2017 | 2019 | ▂▂▂▂▃▃▃▂▂▂ |
| glucose homeostasis | 2017 | 10.14 | 2017 | 2018 | ▂▂▂▂▃▃▂▂▂▂ |
| obese patient | 2019 | 9.30 | 2019 | 2022 | ▂▂▂▂▂▂▃▃▃▃ |
| serum | 2020 | 9.69 | 2020 | 2022 | ▂▂▂▂▂▂▂▃▃▃ |

SUPPLEMENTARY TABLE S3 Top 25 burst references in articles related to the effect of intestinal microbes on obesity. Burst strength indicates the importance of references to the research field. The years between “Begin” and “End” represent the period when the reference was more influential. The thick dark bar represents the year in which the burst reference appeared and the burst duration.

| **References** | **Year** | **Strength** | **Begin** | **End** | **2013 - 2022** |
| --- | --- | --- | --- | --- | --- |
| [Qin JJ, 2012, NATURE, V490, P55, DOI 10.1038/nature11450, DOI](http://dx.doi.org/10.1038%2Fnature11450) | 2012 | 90.81 | 2013 | 2017 | ▃▃▃▃▃▂▂▂▂▂ |
| [Turnbaugh PJ, 2009, NATURE, V457, P480, DOI 10.1038/nature07540, DOI](http://dx.doi.org/10.1038%2Fnature07540) | 2009 | 86.15 | 2013 | 2014 | ▃▃▂▂▂▂▂▂▂▂ |
| [Wu GD, 2011, SCIENCE, V334, P105, DOI 10.1126/science.1208344, DOI](http://dx.doi.org/10.1126%2Fscience.1208344) | 2011 | 77.55 | 2013 | 2016 | ▃▃▃▃▂▂▂▂▂▂ |
| [Qin JJ, 2010, NATURE, V464, P59, DOI 10.1038/nature08821, DOI](http://dx.doi.org/10.1038%2Fnature08821) | 2010 | 72.8 | 2013 | 2015 | ▃▃▃▂▂▂▂▂▂▂ |
| [Caporaso JG, 2010, NAT METHODS, V7, P335, DOI 10.1038/nmeth.f.303, DOI](http://dx.doi.org/10.1038%2Fnmeth.f.303) | 2010 | 71.81 | 2013 | 2015 | ▃▃▃▂▂▂▂▂▂▂ |
| [Schwiertz A, 2010, OBESITY, V18, P190, DOI 10.1038/oby.2009.167, DOI](http://dx.doi.org/10.1038%2Foby.2009.167) | 2010 | 68.86 | 2013 | 2015 | ▃▃▃▂▂▂▂▂▂▂ |
| [Arumugam M, 2011, NATURE, V473, P174, DOI 10.1038/nature09944, DOI](http://dx.doi.org/10.1038%2Fnature09944) | 2011 | 60.45 | 2013 | 2016 | ▃▃▃▃▂▂▂▂▂▂ |
| [Larsen N, 2010, PLOS ONE, V5, P0, DOI 10.1371/journal.pone.0009085, DOI](http://dx.doi.org/10.1371%2Fjournal.pone.0009085) | 2010 | 53.15 | 2013 | 2015 | ▃▃▃▂▂▂▂▂▂▂ |
| [Yatsunenko T, 2012, NATURE, V486, P222, DOI 10.1038/nature11053, DOI](http://dx.doi.org/10.1038%2Fnature11053) | 2012 | 51.79 | 2013 | 2017 | ▃▃▃▃▃▂▂▂▂▂ |
| [Vijay-Kumar M, 2010, SCIENCE, V328, P228, DOI 10.1126/science.1179721, DOI](http://dx.doi.org/10.1126%2Fscience.1179721) | 2010 | 51.18 | 2013 | 2015 | ▃▃▃▂▂▂▂▂▂▂ |
| [Vrieze A, 2012, GASTROENTEROLOGY, V143, P913, DOI 10.1053/j.gastro.2012.06.031, DOI](http://dx.doi.org/10.1053%2Fj.gastro.2012.06.031) | 2012 | 47.11 | 2013 | 2017 | ▃▃▃▃▃▂▂▂▂▂ |
| [Furet JP, 2010, DIABETES, V59, P3049, DOI 10.2337/db10-0253, DOI](http://dx.doi.org/10.2337%2Fdb10-0253) | 2010 | 43.35 | 2013 | 2015 | ▃▃▃▂▂▂▂▂▂▂ |
| [De Filippo C, 2010, P NATL ACAD SCI USA, V107, P14691, DOI 10.1073/pnas.1005963107, DOI](http://dx.doi.org/10.1073%2Fpnas.1005963107) | 2010 | 42.86 | 2013 | 2015 | ▃▃▃▂▂▂▂▂▂▂ |
| [Le Chatelier E, 2013, NATURE, V500, P541, DOI 10.1038/nature12506, DOI](http://dx.doi.org/10.1038%2Fnature12506) | 2013 | 76.08 | 2014 | 2018 | ▂▃▃▃▃▃▂▂▂▂ |
| [Tremaroli V, 2012, NATURE, V489, P242, DOI 10.1038/nature11552, DOI](http://dx.doi.org/10.1038%2Fnature11552) | 2012 | 65.85 | 2014 | 2017 | ▂▃▃▃▃▂▂▂▂▂ |
| [Karlsson FH, 2013, NATURE, V498, P99, DOI 10.1038/nature12198, DOI](http://dx.doi.org/10.1038%2Fnature12198) | 2013 | 49.41 | 2014 | 2018 | ▂▃▃▃▃▃▂▂▂▂ |
| [Everard A, 2013, P NATL ACAD SCI USA, V110, P9066, DOI 10.1073/pnas.1219451110, DOI](http://dx.doi.org/10.1073%2Fpnas.1219451110) | 2013 | 100.42 | 2015 | 2018 | ▂▂▃▃▃▃▂▂▂▂ |
| [David LA, 2014, NATURE, V505, P559, DOI 10.1038/nature12820, DOI](http://dx.doi.org/10.1038%2Fnature12820) | 2014 | 75.51 | 2015 | 2019 | ▂▂▃▃▃▃▃▂▂▂ |
| [Ridaura VK, 2013, SCIENCE, V341, P1079, DOI 10.1126/science.1241214, DOI](http://dx.doi.org/10.1126%2Fscience.1241214) | 2013 | 74.7 | 2015 | 2018 | ▂▂▃▃▃▃▂▂▂▂ |
| [Langille MGI, 2013, NAT BIOTECHNOL, V31, P814, DOI 10.1038/nbt.2676, DOI](http://dx.doi.org/10.1038%2Fnbt.2676) | 2013 | 62.72 | 2016 | 2018 | ▂▂▂▃▃▃▂▂▂▂ |
| [Koh A, 2016, CELL, V165, P1332, DOI 10.1016/j.cell.2016.05.041, DOI](http://dx.doi.org/10.1016%2Fj.cell.2016.05.041) | 2016 | 47.58 | 2019 | 2022 | ▂▂▂▂▂▂▃▃▃▃ |
| [Callahan BJ, 2016, NAT METHODS, V13, P581, DOI 10.1038/NMETH.3869, DOI](http://dx.doi.org/10.1038%2FNMETH.3869) | 2016 | 92.64 | 2020 | 2022 | ▂▂▂▂▂▂▂▃▃▃ |
| [Bolyen E, 2019, NAT BIOTECHNOL, V37, P852, DOI 10.1038/s41587-019-0209-9, DOI](http://dx.doi.org/10.1038%2Fs41587-019-0209-9) | 2019 | 77.15 | 2020 | 2022 | ▂▂▂▂▂▂▂▃▃▃ |
| [Depommier C, 2019, NAT MED, V25, P1096, DOI 10.1038/s41591-019-0495-2, DOI](http://dx.doi.org/10.1038%2Fs41591-019-0495-2) | 2019 | 63.61 | 2020 | 2022 | ▂▂▂▂▂▂▂▃▃▃ |
| [Wang K, 2019, CELL REP, V26, P222, DOI 10.1016/j.celrep.2018.12.028, DOI](http://dx.doi.org/10.1016%2Fj.celrep.2018.12.028) | 2019 | 48.09 | 2020 | 2022 | ▂▂▂▂▂▂▂▃▃▃ |

SUPPLEMENTARY TABLE S4 Top 10 most cited countries/regions related to the effect of intestinal microbes on obesity.

| **Country** | **Citations per article** | **Total Citations** |
| --- | --- | --- |
| Israel | 124.18 | 6085 |
| Belgium | 107.85 | 11648 |
| Sweden | 103.95 | 9148 |
| Austria | 76.65 | 2606 |
| Germany | 59.04 | 10155 |
| Netherlands | 53.61 | 9542 |
| France | 52.71 | 13809 |
| Ireland | 52.44 | 3304 |
| the United States | 52.20 | 87065 |
| Switzerland | 51.27 | 2666 |

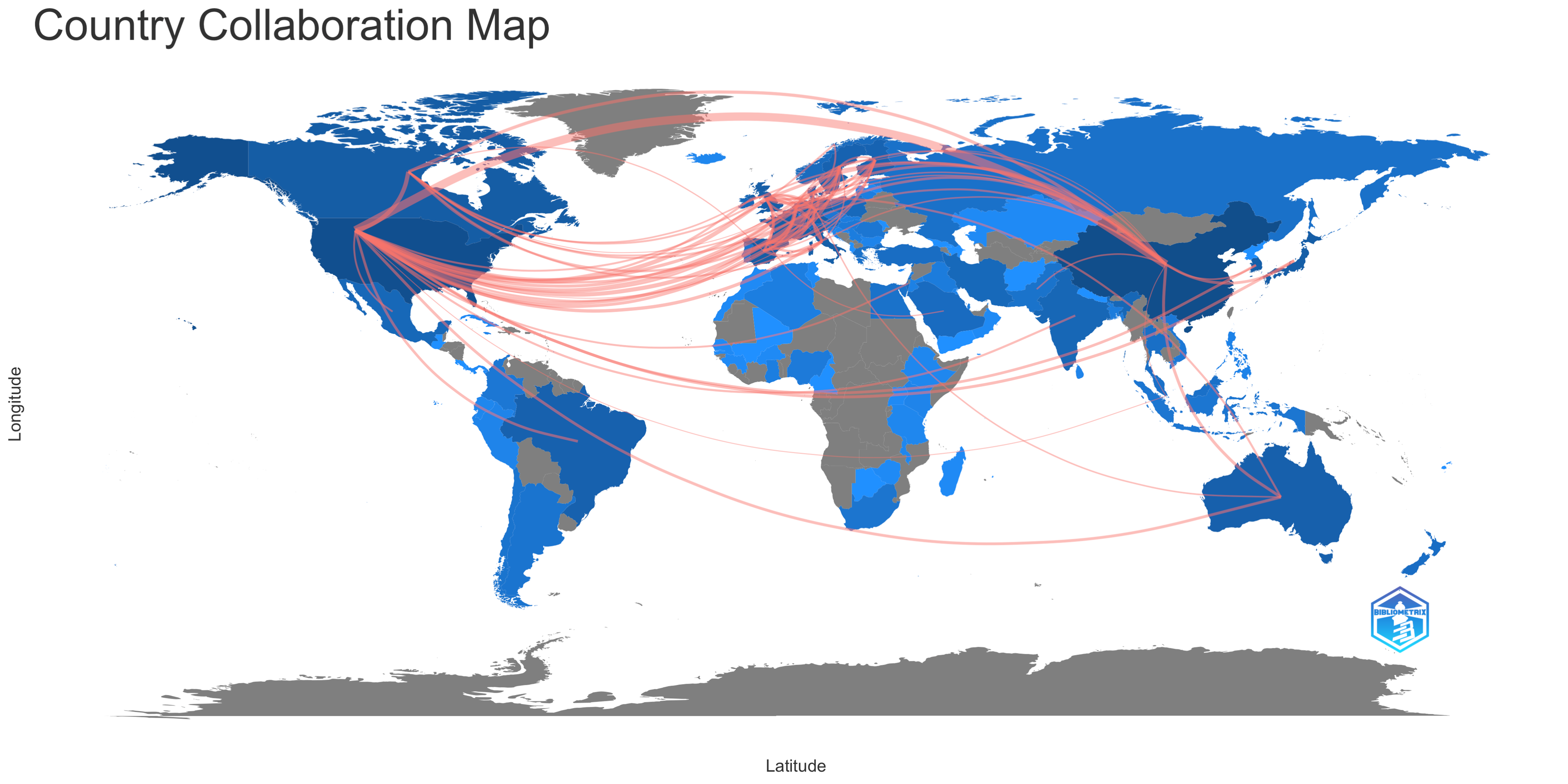

SUPPLEMENTARY FIGURE 1 Country collaboration map. The red line indicates that there was collaboration between two countries. The more connections two countries have, the thicker the line is. The deeper the color of a country is, the more documents are published in this country.

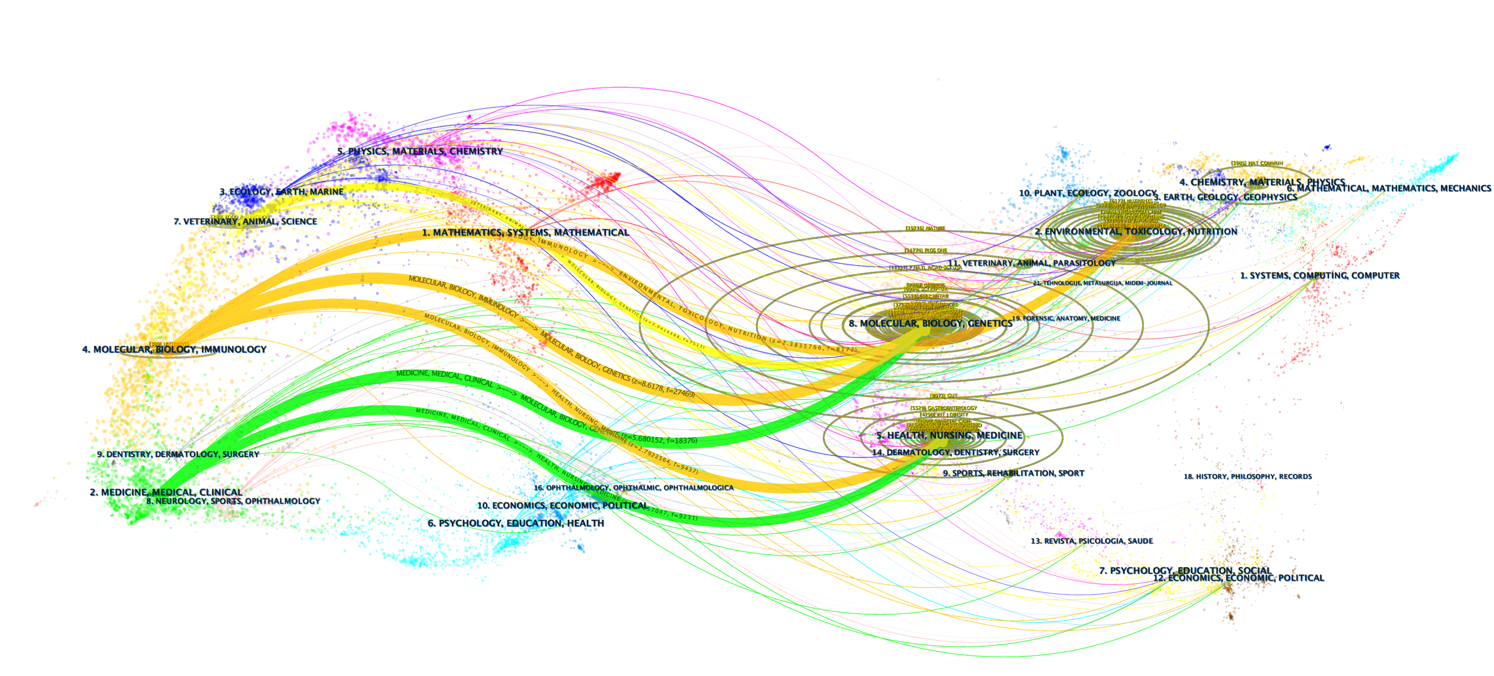

SUPPLEMENTARY FIGURE 2 Dual-map overlay related to the effect of intestinal microbes on obesity.

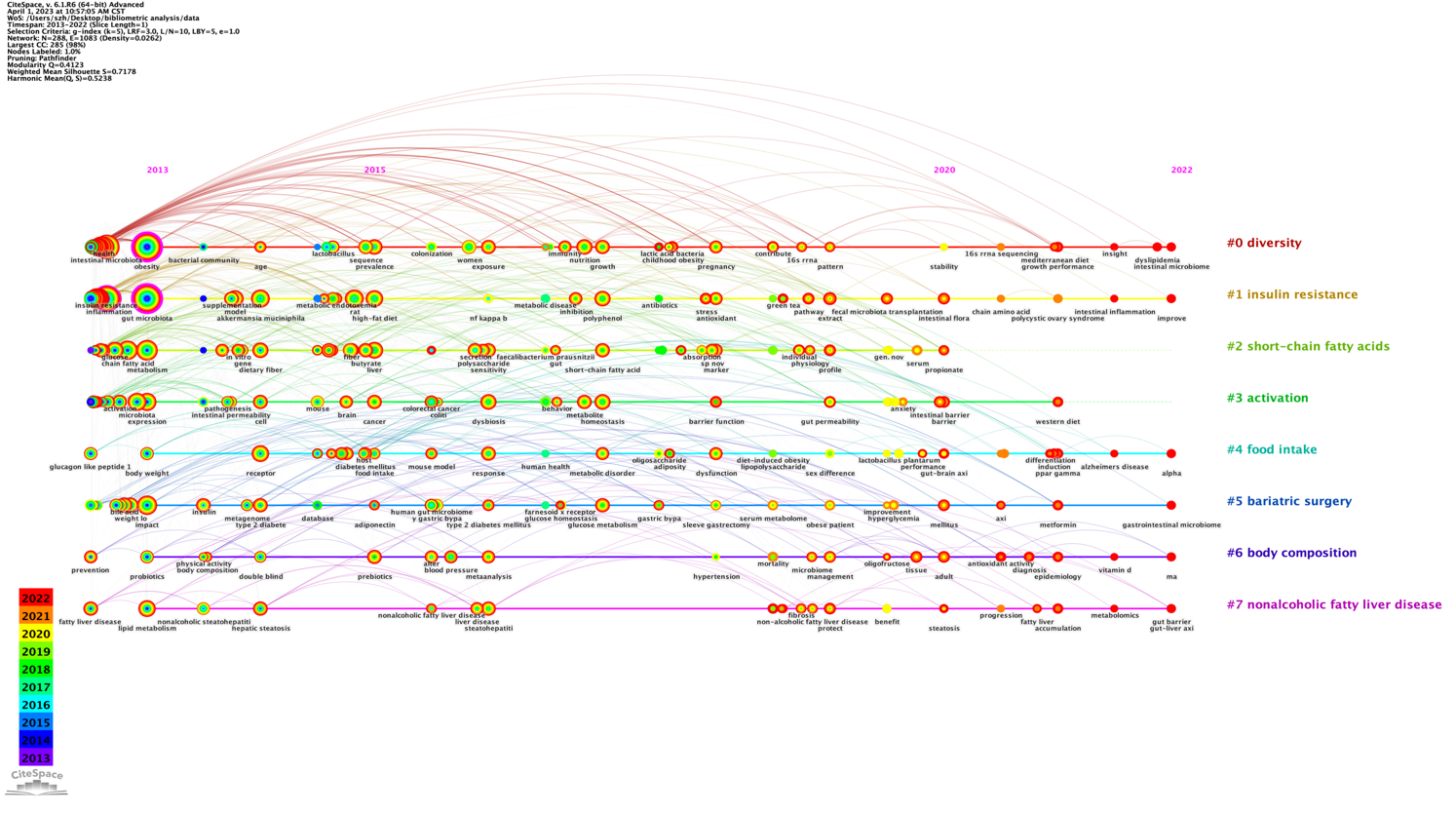

SUPPLEMENTARY FIGURE 3 Cluster timeline view map of keyword analysis for the effect of intestinal microbes on obesity research.

.
